# Psychosocial Health of School-aged Children during the Initial COVID-19 Safer-at-Home School mandates in Florida: A cross-sectional study

**DOI:** 10.1101/2020.11.20.20235812

**Authors:** Sarah McKune, Daniel Acosta, Nick Diaz, Kaitlin Brittain, Diana Joyce-Beaulieu, Anthony T. Maurelli, Eric J. Nelson

## Abstract

**Background:** Given the emerging literature regarding the impacts of lockdown measures on mental health, this study aims to identify risk factors in school-aged children for being at risk for psychosocial disorders during the COVID-19 Safer-at-Home School mandates in Florida

**Methods:** A cross-sectional study was conducted in April 2020 (n=280). Bivariate analysis and logistic and multinomial logistic regression models are used to examine socio-demographic and knowledge, attitude, and practice (KAP) predictors of anxiety, depression, and obsessive-compulsive disorder (OCD).

**Results:** Loss of household income was associated with being at risk for depression [aOR=3.130, 95% CI= (1.41-6.97)], anxiety [aOR=2.531, 95%CI= (1.154-5.551)], and OCD [aOR=2.90, 95%CI= (1.32-6.36)]. Being female was associated with risk for depression [aOR=1.72, 95% CI=(1.02-2.93)], anxiety [aOR=1.75, 95% CI=(1.04-2.97)], and OCD[aOR=1.764, 95%CI= (1.027-3.028)]. Parental practices that are protective against COVID-19 were associated with children being at risk of depression [aOR=1.55, 95% CI= (1.04-2.31)]. Being at a lower school level was risk factor for anxiety and OCD.

**Conclusions:** Efforts to address mental health risk in children, as a result schools should prioritize girls, younger children, and children of families who lose income. Limiting the spread of COVID-19 through school closure may exacerbate the risk of psychosocial disorders in children, thus school administrators should move quickly to target those at greatest risk.

## Background

Social distancing is the primary public health intervention to reduce the spread of SARS-CoV-2, the virus that causes COVID-19. In the Spring of 2020, school closures, limits on the number of individuals allowed at social gatherings, and closing restaurants, bars, and businesses rapidly reduced transmission and flattened the epidemiologic curve in many parts of the US, including Florida. Though children appear to be at less risk of severe illness and mortality associated with COVID-19^1^, these interventions significantly disrupted the lives of children and families – and not equally. Disparities in health care, education, and wealth, which are prevalent across the United States, put minorities and disadvantaged groups at higher risk for negative outcomes associated with the pandemic.

The effects of COVID-19 are far reaching, and scientists urged the research community early in the pandemic to prioritize high quality data on mental health and psychosocial effects of the COVID-19 pandemic ^2,3^. Past evidence has shown significant psychological effects on children during disasters ^4^, and in the context of the COVID-19 pandemic, reports of psychosocial distress in children and adolescents have increased compared to the pre-pandemic baseline ^5,6^. COVID-19 related hardships have also affected psychological wellbeing of children, with greater impact on those who have pre-existing mental illness or who live in households facing larger economic distress^6,7^. Findings from early studies in China found an increase in the prevalence of children reporting symptoms of depression and anxiety during home confinement ^8^. A similar study of the general population in China found that some preventive measures were associated with reporting mental health symptoms ^9^. COVID-19 psychosocial data are limited, especially data representative of racial and ethnically diverse populations of children in the US. Given that minority communities and communities of color in the US have experienced much higher rates of infection and death ^10^, the distribution of the psychological toll of COVID-19, including fear, loss, and trauma, is disproportionately affecting children of these communities ^11,12^. Unequal access to healthcare and educational options during the pandemic, along with any discrimination that comes from being a member of a community with high rates of transmission^5,13^, will only serve to amplify these psychosocial ramifications.

Families have varying degrees of knowledge, and myriad attitudes and practices surrounding COVID-19. If/how these knowledge, attitudes, and practices (KAP) are associated with risk of poor psychosocial outcomes in children is unknown. KAP studies, often used to develop appropriate public health interventions, are built on the well-established Theory of Planned Behavior and assume a linear relationship, culminating in behaviors that improve health ^14–16^. Given that the situation with COVID-19 has had high levels of uncertainty, there might be a relationship with KAP and psychosocial distress.

Understanding the effects of COVID-19 on the psychosocial health of school-aged children and identifying those groups of children most at risk is necessary for the development of appropriate, targeted mental health interventions. The aim of this study is to describe the psychosocial health of a population of school-aged children during the early phase of the COVID-19 pandemic. We examine parental KAP related to COVID-19 by race/ethnicity. We then examine these, and other socio-demographic risk factors associated with poor psychosocial health in school-aged children. This information will assist school administrators, public health practitioners, and policy makers in designing evidence-based, targeted interventions to address the psychosocial needs of the school-aged population during the ongoing COVID-19 pandemic. This study examines social groups (sex, age, race, and ethnicity) and COVID-19 related KAP as risk factors for indicators of poor psychosocial outcomes, including anxiety, depression, and obsessive compulsive disorder (OCD), among school-aged (K-12) children in Florida at the beginning of the pandemic.

## Methods

### Study Design

A cross-sectional study was conducted in April 2020 to describe the effects of COVID-19 on school-aged children and their parents. We present results from an online survey that collected information on socio-demographics, clinical risk factors, parental COVID-19 related KAP, and indicators of psychosocial risk in children. The survey was coupled with the collection of oropharyngeal swabs (PCR for viral detection) and finger sticks (ELISA for antibodies) from student participants at a drive-thru testing site. Laboratory data associated with this study will be published independently and are not included herein.

Data were collected at a public, K-12 school in Florida. The school is mandated to have a student body that reflects the demographics of the State of Florida. Students are characterized by gender, race/ethnicity, family income, and academic needs. Parents/guardians of all current students (N=1178) were invited by email to have their child participate in the study. Those who were interested were able to consent/assent to participation online via a HIPAA compliant interface connected to RedCap (Vanderbilt University). Parental consent was required for all children under the age of 18; and, all children over the age of eight assented for themselves. All data were deidentified prior to analysis and were stored on secured servers to ensure the protection of participants (RedCap). The study was approved by the University of Florida Institutional Review Board, protocol IRB202001345

### Variables and statistical analyses

The demographics section of the survey included questions about child’s age, sex, race, ethnicity, and the loss of household income. Other risk factors evaluated in the survey included, clinical symptoms and parental occupation. The COVID-19 related KAP section consisted of fourteen knowledge, eight attitude, and five practice questions. Knowledge scores were created by assigning one (1) to each correctly answered knowledge question about COVID-19 (transmission, prevention, and/or general information) and summing them for a total possible score of 14. Similarly, attitudes and practices were categorized as protective and unprotective, with protective attitudes and practices coded as one (1) and all others as zero (0), and summed, for respective possible scores of 8 and 5.

The questions used to measure psychosocial risk evaluate three internalizing disorders: depression, anxiety, and OCD. These emotional and behavioral disorders were chosen because of their critical importance and generalizability to the pediatric COVID-19 literature^6,8,17^. Importantly, externalizing disorders are not evaluated in this study. Each psychosocial disorder (depression, anxiety, and OCD) was evaluated using a set of categorical questions (5-point Likert scale). All questions used age-appropriate language and response options (e.g. 1-Never, 2-A Little, 3-Sometimes, 4-A Lot, or 5-Always/Constantly). Within each group of questions, if a participant’s responses were all 1s or 2s, the child was categorized Not at Risk; everyone else was considered At Risk and further categorized as *High, Medium, or Low Risk*, using the frequency of their highest response options (see Likert scale above). Participants who answered “a lot” (4) to two or more questions in a group or “always/constantly” (5) to any one of the questions in the group were considered *High Risk*; participants who answered “a lot” (4) only once, with all other questions at a 3 or below, were considered *Medium Risk*; and any participant who answered “sometimes” (3) to one or more question was considered *Low Risk*. Each group of questions was designed by a team of psychologists working with the research team to identify students at risk of developing one of the specific internalized disorders mentioned above. The six primary outcome variables in this study are *At Risk (Using No Risk as reference (REF))* and *High Risk (REF No Risk)* of anxiety, depression, and OCD. A summary outcome variable, *Any Risk* indicates if a child presents as *At Risk* for any of the three psychosocial outcomes assessed here.

Bivariate analysis of all outcome variables was conducted to test for association with knowledge score, attitude score, and practice score, demographic variables, and parental occupation. An additional bivariate analysis of each individual KAP question was used to identify association between any KAP item (question) and race/ethnicity using Fisher’s Exact Test and logistic regression. Further analysis was conducted using logistic and multinomial logistic regression models to examine predictors of *Any Risk (REF No Risk)* and *High Risk (REF No Risk)*, respectively, for each depression, anxiety, and OCD. The covariates included in the models were race/ethnicity, sex, school level, household loss of income during the pandemic, parent working in a medical setting, knowledge score, attitude score, and practice score. Survey data were collected in RedCap and analyzed in R Software (version 4.0.0) and SPSS (version 26).

## Results

A total of 280 students were enrolled out of student body of 1178. Table 1 presents the demographic characteristic of the sample and the adjusted odds ratio (aOR) for being at *Any Risk* for one or more of three internalizing syndromes considered in this study (depression, anxiety, or OCD). The study population was nearly 2/3 White, a fifth Hispanic (regardless of race), and 9% Black. Participants were 48% male, and distributed across high school (40%), middle school (29%) and primary school (31%).

**Table 1:**
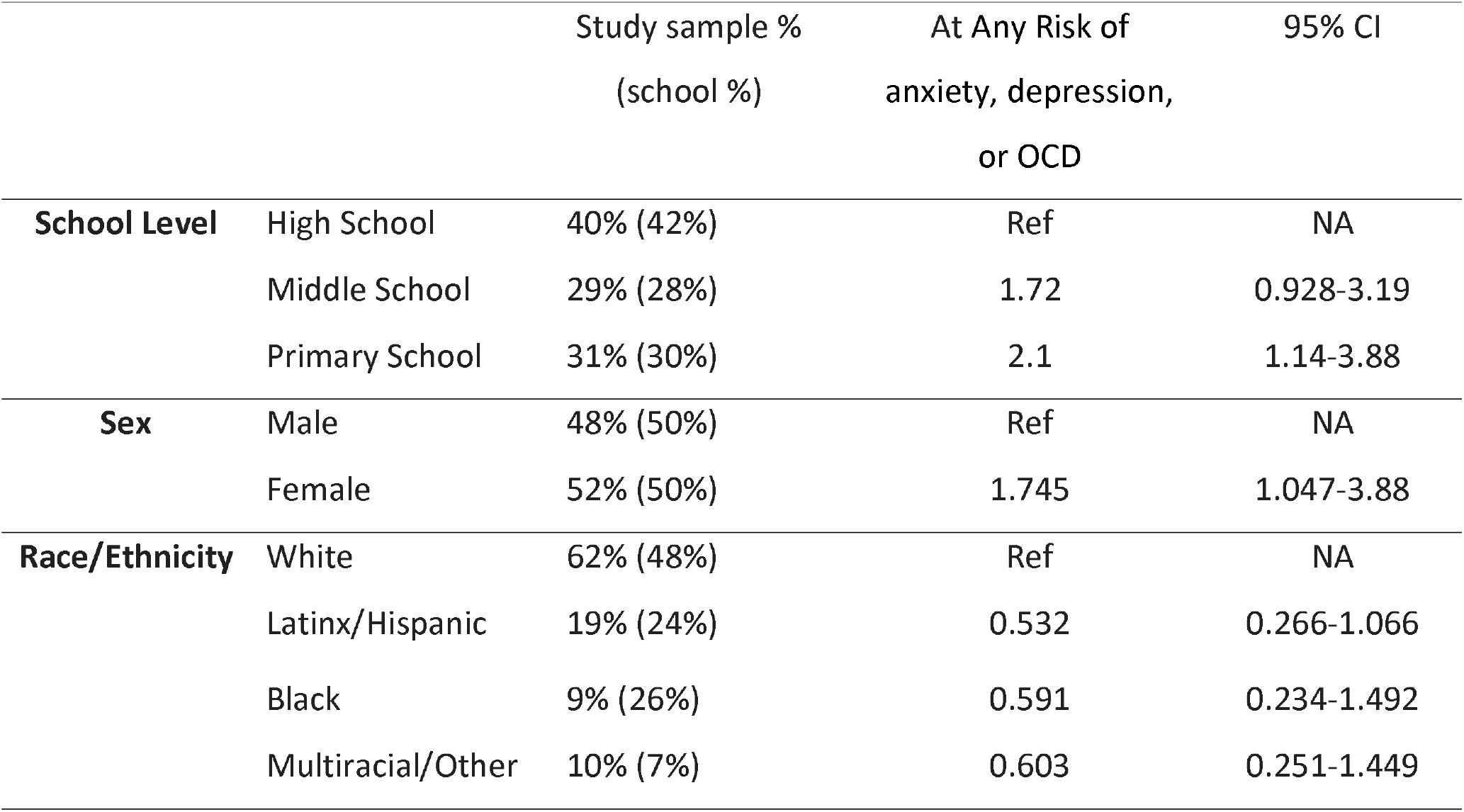
Demographic characteristics of the study population and Adjusted Odds Ratio (aOR) for being at Any Risk of anxiety, depression, or OCD (n=280)

|  |  | Study sample %<br>(school %) | At Any Risk of<br>anxiety, depression,<br>or OCD | 95% CI |
| --- | --- | --- | --- | --- |
| **School Level** | High School | 40% (42%) | Ref | NA |
|  | Middle School | 29% (28%) | 1.72 | 0.928-3.19 |
|  | Primary School | 31% (30%) | 2.1 | 1.14-3.88 |
| **Sex** | Male | 48% (50%) | Ref | NA |
|  | Female | 52% (50%) | 1.745 | 1.047-3.88 |
| **Race/Ethnicity** | White | 62% (48%) | Ref | NA |
|  | Latinx/Hispanic | 19% (24%) | 0.532 | 0.266-1.066 |
|  | Black | 9% (26%) | 0.591 | 0.234-1.492 |
|  | Multiracial/Other | 10% (7%) | 0.603 | 0.251-1.449 |

Parental knowledge about COVID-19 was high, with 31.4% answering all questions correctly, and only 13.6% answering three or more questions incorrectly (Table 2). Attitudes had similar results, with most parents expressing agreement with protective attitudes. Preventative practices were also high, with at least 90% of respondents reporting increased hand washing, avoiding physical contact with those outside their home, and adhering to social distancing guidelines (Table 3).

**Table 2:**
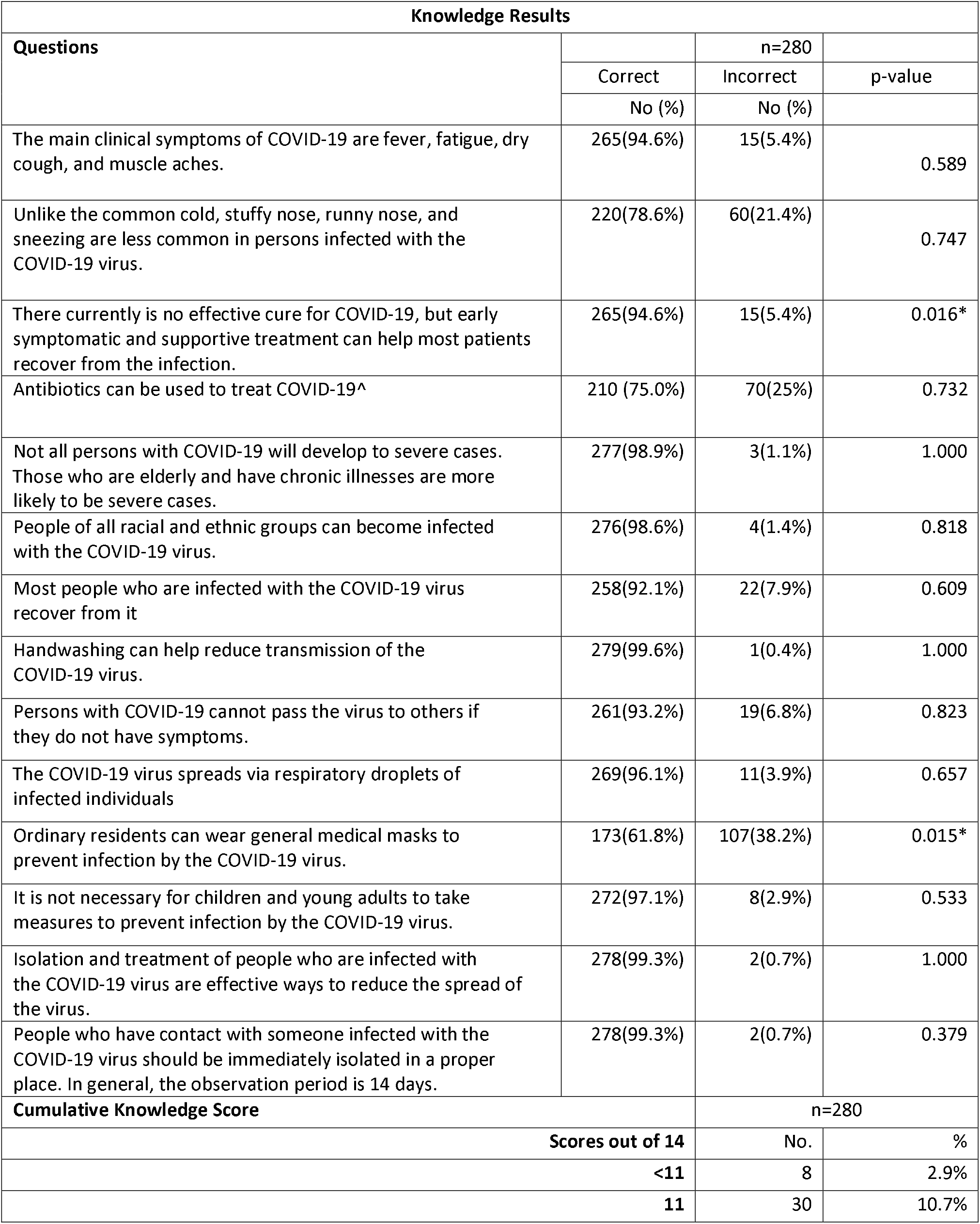
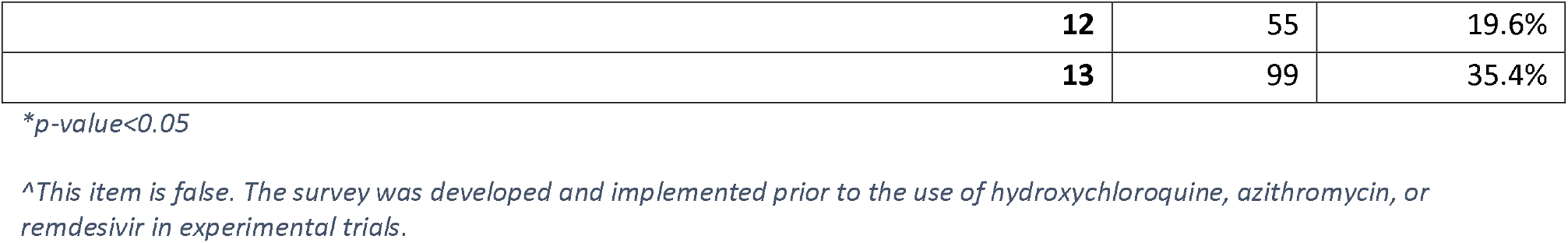
Knowledge Results. P-values are for Fisher’s Exact Test comparing knowledge answers and race/ethnicity.

**Table 3:**
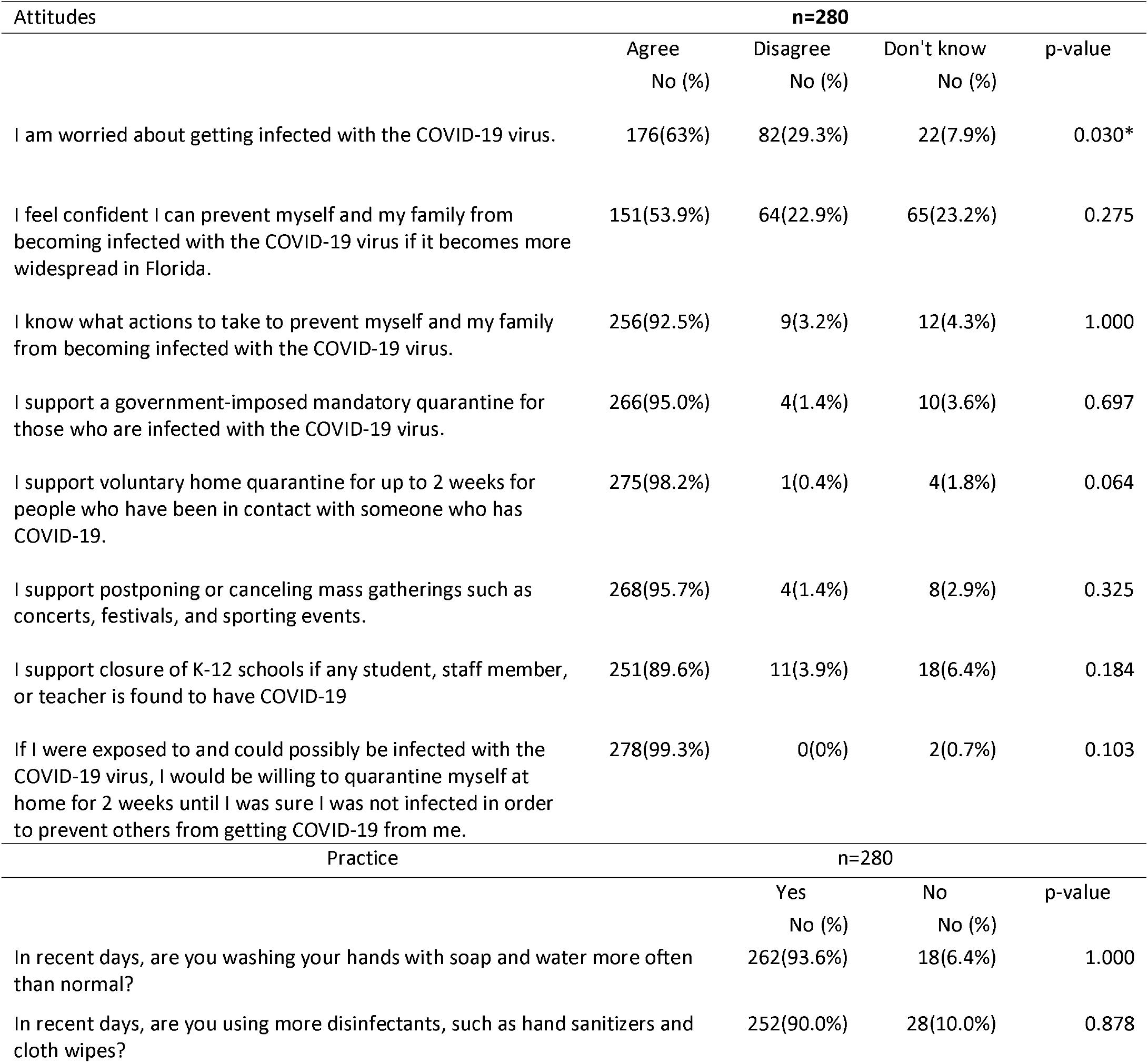
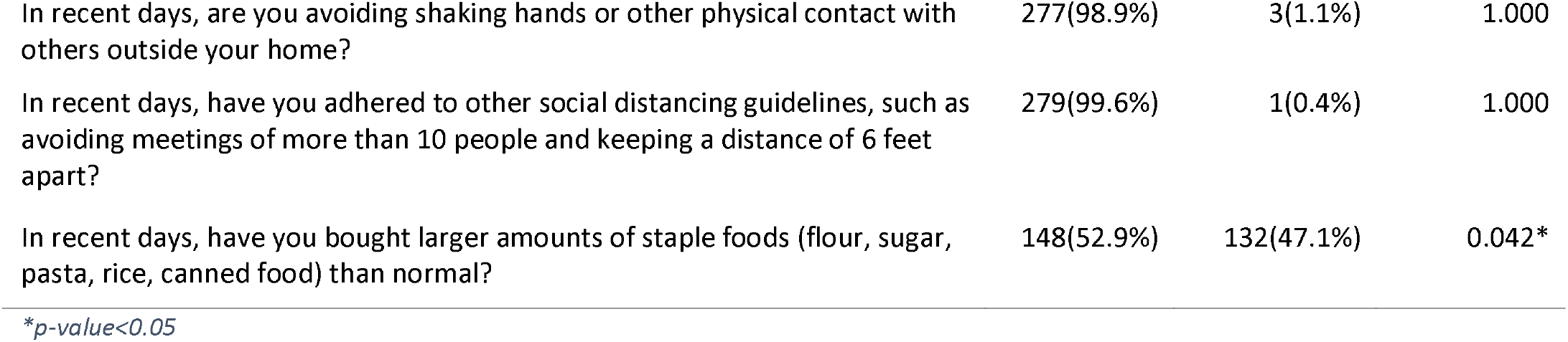
Attitudes and Practices Results. P-values are for Fisher’s Exact Test comparing protective and un-protective attitudes and practices with race/ethnicity.

| Attitudes | n=280 |  |  | p-value |
| --- | --- | --- | --- | --- |
|  | Agree<br>No (%) | Disagree<br>No (%) | Don't know<br>No (%) |  |
| I am worried about getting infected with the COVID-19 virus. | 176(63%) | 82(29.3%) | 22(7.9%) | 0.030* |
| I feel confident I can prevent myself and my family from becoming infected with the COVID-19 virus if it becomes more widespread in Florida. | 151(53.9%) | 64(22.9%) | 65(23.2%) | 0.275 |
| I know what actions to take to prevent myself and my family from becoming infected with the COVID-19 virus. | 256(92.5%) | 9(3.2%) | 12(4.3%) | 1.000 |
| I support a government-imposed mandatory quarantine for those who are infected with the COVID-19 virus. | 266(95.0%) | 4(1.4%) | 10(3.6%) | 0.697 |
| I support voluntary home quarantine for up to 2 weeks for people who have been in contact with someone who has COVID-19. | 275(98.2%) | 1(0.4%) | 4(1.8%) | 0.064 |
| I support postponing or canceling mass gatherings such as concerts, festivals, and sporting events. | 268(95.7%) | 4(1.4%) | 8(2.9%) | 0.325 |
| I support closure of K-12 schools if any student, staff member, or teacher is found to have COVID-19 | 251(89.6%) | 11(3.9%) | 18(6.4%) | 0.184 |
| If I were exposed to and could possibly be infected with the COVID-19 virus, I would be willing to quarantine myself at home for 2 weeks until I was sure I was not infected in order to prevent others from getting COVID-19 from me. | 278(99.3%) | 0(0%) | 2(0.7%) | 0.103 |
| Practice | n=280 |  |  | p-value |
|  | Yes<br>No (%) | No<br>No (%) |  |  |
| In recent days, are you washing your hands with soap and water more often than normal? | 262(93.6%) | 18(6.4%) |  | 1.000 |
| In recent days, are you using more disinfectants, such as hand sanitizers and cloth wipes? | 252(90.0%) | 28(10.0%) |  | 0.878 |

|  |  |  |  |
| --- | --- | --- | --- |
| In recent days, are you avoiding shaking hands or other physical contact with others outside your home? | 277(98.9%) | 3(1.1%) | 1.000 |
| In recent days, have you adhered to other social distancing guidelines, such as avoiding meetings of more than 10 people and keeping a distance of 6 feet apart? | 279(99.6%) | 1(0.4%) | 1.000 |
| In recent days, have you bought larger amounts of staple foods (flour, sugar, pasta, rice, canned food) than normal? | 148(52.9%) | 132(47.1%) | 0.042 * |

\*p-value<0.05

There was a statistically significant association between race/ethnicity and knowledge questions regarding the existence of an effective cure of the virus, where Multiracial/Other respondents had a higher rate of incorrect responses compared to White respondents. The use of masks as a protective factor against COVID-19 was also significant, with White respondents answering that masks do not protect against the virus at a higher proportion than African Americans and Hispanics (Table 2).

There were race/ethnic differences in who was worried about getting infected by the virus and, correspondingly, with the knowledge question about mask usage; White respondents were less worried overall about getting infected with the virus compared to African Americans and Hispanics (Table 3). There were no significant differences between Multiracial/Other and White respondents in mask usage or being worried about infection with the virus. Among those practices assessed, no significant difference by race/ethnicity was found for social distancing, handwashing, or general hygiene; however, Hispanic respondents were less likely to report purchasing larger amounts of staple foods than normal.

Loss of household income was significantly associated with students being At Risk of depression [aOR=3.130, 95% CI= (1.41-6.97)], anxiety [aOR=2.531, 95%CI= (1.154-5.551)], and OCD [aOR=2.90, 95%CI= (1.32-6.36)], see Table 4. Statistical results also indicate a marginally significant association (where p< 0.1) between loss of household income and a student being at *High Risk* for depression [p value 0.05, aOR=3.74, 95% CI=(1.00-14.01)] and OCD [p value 0.069, aOR=3.19, 95% CI=(0.91-11.12)], see Table 4. Being female was significantly associated with being At Risk for depression [aOR=1.72, 95% CI=(1.02-2.93)], anxiety [aOR=1.75, 95% CI=(1.04- 2.97)], and OCD[aOR=1.764, 95%CI= (1.027-3.028)]. Girls may also be more likely to present as High Risk for depression [p value=0.057, aOR=2.93, 95% CI= (0.970-8.84), Table 3] and OCD [p value= 0.050 aOR=2.58, 95%CI= (0.99-6.68), table 4], as a marginally significant association was identified.

**Table 4.** Adjusted Odds Ratios Children At Risk and at High Risk for depression, anxiety, and OCD, by race/ethnicity, sex, school level, household loss of income, parental employment, and COVID-19 KAP.

| Covariates | Anxiety |  | Depression |  | OCD |  |
| --- | --- | --- | --- | --- | --- | --- |
|  | aOR At Risk (95% C.I.)<sup>1</sup> | aOR High Risk (95% C.I.)<sup>2</sup> | aOR At Risk (95% C.I.)<sup>1</sup> | aOR High Risk (95% C.I.)<sup>2</sup> | aOR At Risk (95% C.I.)<sup>1</sup> | aOR High Risk (95% C.I.)<sup>2</sup> |
| **Race/Ethnicity** |  |  |  |  |  |  |
| White | Ref (NA) | Ref (NA) | Ref (NA) | Ref (NA) | Ref (NA) | Ref (NA) |
| Black | 0.758(0.29-1.97) | 0.34(0.04-2.96) | 0.97(0.39 -2.48) | 0.74(0.082-6.78) | 1.01(0.39-2.66) | 2.17(0.57(8.37) |
| Hispanic | 0.589(0.28-1.22) | 0.82(0.26-2.64) | 0.68(0.33-1.42) | 1.83(0.56-5.99) | 1.02(0.49-2.08) | 0.79(0.22-2.82) |
| Multiracial | 1.034(0.43-2.49) | 1.47(0.34-6.36) | 0.78 (0.31-1.96) | 2.53(0.49-11.41) | 1.41(0.58-3.43) | 1.55(0.35-6.87) |
| **Sex** |  |  |  |  |  |  |
| Male | Ref (NA) | Ref (NA) | Ref (NA) | Ref (NA) | Ref (NA) | Ref (NA) |
| Female | 1.753(1.04-2.97) | 1.93(0.76-4.88) | 1.72 (1.02-2.93) | 2.93(0.970-8.84) | 1.76(1.03-3.03) | 2.58(0.99-6.68) |
| **School Level** |  |  |  |  |  |  |
| High School | Ref (NA) | Ref (NA) | Ref (NA) | Ref (NA) | Ref (NA) | Ref (NA) |
| Middle School | 1.82(0.95-3.56) | 1.26(0.37-4.33) | 1.67(0.88-3.18) | 0.83(0.23-2.99) | 1.16(0.59-2.26) | 1.46(0.44-4.87) |
| Primary School | 2.32(1.23-4.36) | 2.94(1.04-8.30) | 1.63 (0.88-3.07) | 1.03(0.33-3.26) | 2.01(1.07-3.77) | 3.10(1.05-9.12) |
| **Household loss of income due to COVID-19** |  |  |  |  |  |  |
| No | Ref (NA) | Ref (NA) | Ref (NA) | Ref (NA) | Ref (NA) | Ref (NA) |
| Yes | 2.53(1.15-5.55) | 1.17(0.23-5.8) | 3.13(1.41-6.97) | 3.74(1.00-14.01) | 2.90(1.32-6.36) | 3.19(0.91-11.12) |
| **Parent working on frontline** |  |  |  |  |  |  |
| No | Ref (NA) | NA | Ref (NA) | Ref (NA) | Ref | NA |
| Yes | 0.66(0.37-1.18) | 0.41(0.13-1.25) | 0.59(0.33-1.07) | 0.43(0.13-1.45) | 0.59(0.32-1.09) | 0.42(0.14-1.31) |
| **KAP Scores** |  |  |  |  |  |  |
| Knowledge Score | 1.05(0.83-1.32) | 0.96(0.64-1.43) | 0.96(0.77-1.21) | 1.11(0.69-1.76) | 0.89(0.77-1.25) | 0.72(0.50-1.04) |
| Attitude Score | 0.97(0.75-1.25) | 1.82(0.97-3.43) | 0.90(0.71-1.16) | 1.51(0.79-2.89) | 0.25(0.89-1.56) | 1.46(0.87-2.44) |
| Practice Score | 1.08(0.75-1.58) | 1.19(0.57-2.52) | 1.55(1.04-2.31) | 1.34(0.63-2.86) | 0.42(0.79-1.74) | 1.13(0.57-2.22) |

<sup>1</sup> Binary Logistic Regression was used to model At Risk, using No Risk as the reference category
<sup>2</sup> Multinomial Logistic Regression was used to model High Risk using No Risk as the reference category

School level was significantly associated with being At Risk and *High Risk* of both anxiety and OCD (Table 4). Those in primary school and middle school are more likely to be At Risk than students in high school, and those in primary school being more likely to be at *High Risk*. A family’s COVID-19 Practice score was significantly associated with a child being At Risk for depression [aOR=1.55, 95% CI= (1.04-2.31), Table 4]; those families who followed stricter protective practices were more likely to have a child who presented as *At Risk* for depression. A family’s COVID-19 Attitude score was marginally significant in the multinomial regression model for *High Risk* of anxiety [p value=0.062, aOR 1.82, 95% CI (0.971-3.43)]. Inversely, increased parental COVID-19 Knowledge was protective against children presenting as High Risk for OCD. Parents working in a frontline medical setting was protective against children being At Risk for depression [p value=0.084, aOR=0.59, 95% CI= (0.33-1.07)] or OCD [p value= 0.091 aOR=0.592, 95%CI= (0.322-1.088)].

## Discussion

A number of items present in this analysis are risk factors for psychosocial distress among school-aged children. These include a loss of income in their household, being female, and being in primary school. In addition, families who report more protective practices were more likely to have children who present as *At Risk* for depression, and families who indicate attitudes consistent with protection from COVID-19 are more likely to have children who present as high risk for anxiety. These results merit further discussion.

Loss of household income was a clear indicator of mental distress across the sample population, which is congruent with other studies, suggesting that the economic impacts of the lockdown are an important trigger for mental distress in children.^3,7,9^. Economic hardship alone might be associated with increased risk of socioemotional problems in children, exacerbated by their parents’ response to the situation ^18^. This may also be a partial explanation for why having a parent working on the medical frontline was marginally significant as a protective factor for depression and OCD, as those may be households that are economically advantaged (doctors, nurses), and did not lose income compared to others.

Sex was also an important risk factor, as girls were more likely to be at risk for depression, anxiety, and OCD. This finding echoes results from post-disaster related studies, where being female was associated with increased psychosocial risk ^19^On the other hand, this result might be related to reporting, as girls are more likely than boys to report their risk and/or clinical symptoms of depression ^20–22^. The results also suggest that children in primary school are more likely to present as at high risk for anxiety and OCD, making them priority groups for interventions at schools.

Reporting protective practices was associated with children presenting as At Risk for depression. Distancing and isolation can increase stress, which can aggravate feelings of loneliness and impact long term health^3^.. Protective attitudes in parents was also associated with children presenting as high risk for anxiety. Higher practice scores were associated with hand washing, avoiding physical contact, and adhering to social distancing guidelines, while higher attitudes scores are related to supporting closures and limiting social gatherings. Even though these attitudes and practices are crucial to slow the spread of viral diseases, to continue implementing them might increase risk of psychosocial distress.

Thus, the pandemic paradox arises: do we limit the spread of disease at the cost of higher risk of psychosocial distress, or do we risk overwhelming the health system and witness a spike in deaths, as we seek to protect children from risk of psychosocial distress? There is no simple answer, as both options have important negative impacts and implications. Extended quarantine can expose children to domestic violence ^23–25^, as well as aggravating loss of household income to those who are unable to hire private childcare, thus are unable to return to work. On the other hand, reopening schools could expose children to deaths of teachers, staff, and fellow students, which would only exacerbate negative psychosocial impacts. Additionally, there are important disparities that will influence which children become infected: a recent study of pediatric cases showed that 51% of the children infected came from low-income communities, while only 2% came from high-income communities^26^. Yonker et al. also suggest that children from lower income settings pose a larger threat to their families as household size may be larger with multi-generational co-habitation and higher household density ^26^.

## Conclusion

Many studies aim to predict how lockdown measures will flatten the pandemic curve, but few studies focus on the psychosocial impacts of these interventions on children and their families. As public health experts focus on reducing the spread of COVID-19 infections, it is imperative that they also focus on addressing the psychosocial needs of children. Additional research is needed to better understand and address the impacts of the pandemic and its societal responses on children, but doing so must be inclusive of vulnerable populations, including those whose households have lost income. Future research must include economic status and ensure diversity and inclusion of minorities in order to understand how the impacts affect vulnerable groups differently. Given that loss of household income was clearly an important risk factor for depression, anxiety, and OCD, understanding a household’s baseline economic status becomes important in considering vulnerability and possible mitigating conditions. These findings have important implications for policy makers as they negotiate the continuation of funding for unemployment benefits and other safety nets for those economically disadvantaged, as losing these could increase the number of children at risk of psychosocial distress. There should be a national, coordinated effort to continue collecting psychosocial data on children, as well as to design strategies and coping mechanisms for these children as societies across the country strive to balance the risks of COVID-19 infection and the risk of lockdown on psychosocial wellbeing. Previous disasters have shown that adverse psychological effects are not only present during the event, but also remain long after the incident ^19^. Efforts to address mental health cannot wait until the pandemic is over, neither for school age population as the present study suggests, nor for adults, as other studies indicate^7,9,11,27–29^. At the very minimum, high risk groups must be identified across the US. These data contribute to evidence that lay bare the urgent need for primary and secondary school administrators, in collaboration with public health practitioners and medical professionals, to roll out targeted and group interventions as early as possible to respond to the emotional and psychosocial needs of children after a disaster occurs.

## Limitations

Data were not collected on household income; only loss of income due to COVID-19 was captured, thus limiting interpretation of some of these analyses. The survey instrument was designed in March 2020, when the use of masks was not widely recommended^30^ and there were many uncertainties about the virus and its spread. As a result, only one question about mask usage was included in the knowledge section of the questionnaire. The survey was completed in mid-April when schools had already moved to remote learning and prevention campaigns were already in effect, which could explain the high rate at which knowledge questions were answered correctly.

## Data Availability

All data will be de-identified and made publicly available 12 months after the end of the cohort study.

## List of abbreviations

aOR: Adjusted odds ratio
CI: Confidence Interval
HIPPA: Health Insurance Portability and Accountability Act
KAP: Knowledge, attitudes, and practices
OCD: Obsessive Compulsive Disorder

## Declarations

### Ethics approval and consent to participate

The study was approved by the University of Florida Institutional Review Board, protocol IRB202001345

### Consent for publication

Not applicable

## Authors’ Contributions

SM designed and supervised the study, supervised the analyses, and led the writing. DA completed the analyses and significantly contributed to the writing. ND assisted with the analyses and assisted with the writing. KB assisted with the analyses and assisted with the writing. DJB assisted with the design, with the analyses, and assisted with the writing. ATM designed and supervised the study, assisted with the writing. EJN designed and supervised the study, contributed to the writing

## Acknowledgements

A special thanks to the school personnel and staff; the medical, public health, and nursing student volunteers; Drs. Tara Sabo-Atwood, John Lednicky, and the entire research testing lab staff in Environmental and Global Health; and Dr. Glenn Morris, and Dr. Michael Lauzardo for their support and guidance.

